# Variation in influenza vaccine assessment, receipt, and refusal by the concentration of Medicare Advantage enrollees in U.S. nursing homes

**DOI:** 10.1101/2021.10.06.21264537

**Authors:** Patience Moyo, Elliott Bosco, Barbara H. Bardenheier, Maricruz Rivera-Hernandez, Robertus van Aalst, Ayman Chit, Stefan Gravenstein, Andrew R. Zullo

## Abstract

**Background:** More older adults enrolled in Medicare Advantage (MA) are entering nursing homes (NHs), and MA concentration could affect vaccination rates through shifts in resident characteristics and/or payer related influences on preventive services use. We investigated whether rates of influenza vaccination and refusal differ across NHs with varying concentrations of MA-enrolled residents.

**Methods:** We analyzed 2014-2015 Medicare enrollment data and Minimum Data Set clinical assessments linked to NH-level characteristics, star ratings, and county-level MA penetration rates. The independent variable was the percentage of residents enrolled in MA at admission and categorized into three groups. We examined three NH-level outcomes: percentage of residents assessed and appropriately provided the influenza vaccine, receiving influenza vaccine, and refusing influenza vaccine.

**Results:** There were 936,513 long-stay residents in 12,384 NHs. Categories for the prevalence of MA enrollment in NHs were low (0% to 3.3%; n=4131 NHs), moderate (3.4% to 18.6%; n=4127 NHs) and high (>18.6%; n=4126 NHs). Adjusting for covariates, influenza vaccination rates among long-stay residents were higher in NHs with moderate (1.7%, *P*<0.0001), or high (3.1%, *P*<0.0001) MA versus the lowest prevalence of MA. Influenza vaccine refusal was lower in NHs with moderate (−3.1%, *P*<0.0001), or high (−4.6% *P*<0.0001) MA compared with NHs with the lowest prevalence of MA. Among 753,616 short-stay residents in 12,205 NHs, there was no association between MA concentration and influenza vaccination receipt but vaccine refusal was greater in NHs with higher MA prevalence (high or moderate vs. low MA: 5.2%, *P*<0.0001).

**Conclusion:** The relationship between MA concentration and influenza vaccination measures varied among post-acute and long-term NH residents. As MA takes a larger role in the Medicare program, and more MA beneficiaries enter NHs, there is need to consider how managed care can be leveraged to improve the delivery of preventive services such as influenza vaccinations in NH settings.

## BACKGROUND

Despite Medicare coverage with no out-of-pocket cost to the beneficiary, and CMS vaccination requirements for nursing homes (NHs), influenza vaccination coverage in NHs remains suboptimal.[1, 2] At 73.1%, during the 2018-2019 influenza season, vaccination coverage among adults living in NHs fell short of the Healthy People 2020 goal of 90%.[3] The coronavirus disease 2019 (COVID-19) pandemic has not only placed a spotlight on the vulnerability of NH residents to morbidity and mortality from respiratory infections, but it also has emphasized racial and socioeconomic inequities in patient care and outcomes.[4, 5] Influenza vaccination can reduce influenza severity and prevent hospitalizations to help avoid “twindemic” effects of influenza and COVID-19 co-circulation on the healthcare system and society.[6, 7] The widespread provision and acceptance of influenza vaccines is central to any effective strategy to mitigate the spread of influenza infection in NHs. Thus, efforts to promote vaccine uptake in NHs require careful consideration of several factors, including resident profiles, facility attributes, and type of health insurance coverage.

Enrollment in Medicare Advantage (MA) is growing and is projected to increase to 51% of all Medicare beneficiaries in 2030 from 34% in 2019.[8] Simultaneously, there is rising enrollment of racial and ethnic minorities in MA, and the health profiles of MA and Traditional Medicare (TM) enrollees are increasingly similar over time.[9-14] The historical selection of healthier beneficiaries into MA has diminished (if not reversed) because Medicare implemented changes reducing incentives for MA plans to select enrollees with more favorable risk profiles.[15] MA is distinct from TM in that the Centers for Medicare and Medicaid Services (CMS) pay private health insurance plans on a fixed capitated fee to provide health benefits for Medicare beneficiaries. Features of MA plans such as their payment models, coordinated care, and outreach programs urging high-risk members to get vaccinated may encourage screening and preventive care use to prevent costly medical services.[16]

Research among community-dwelling beneficiaries has found higher rates of preventive services use (e.g., mammography screening, annual influenza vaccinations, and cholesterol testing) in MA compared with TM.[17-19] However, these studies did not consider NH populations and rely on data from more than a decade ago. MA plans and the characteristics of their enrollees have changed over time, as has the population of individuals receiving care in NHs.[20, 21] The composition of MA beneficiaries in a NH and its relationship with the proportion of residents vaccinated has not been characterized. Yet, this is an important lens through which to examine and address gaps in influenza vaccination coverage in NHs, especially as MA plans may be selectively contracting with NHs, such as those that are larger and are part of a chain.[22]

Furthermore, the direction of relationship between MA concentration and NH vaccination rates is uncertain. The shifts in the composition of MA enrollees and the payment model that incentivizes preventive services potentially present opposing possibilities for vaccination rates in NHs. A ‘financial incentives’ hypothesis may suggest that an increased proportion of NH residents enrolled in MA produces higher influenza vaccination rates; participating plans that promote the health of their enrollees in NHs may emphasize preventive services such as influenza vaccinations.[23, 24] Another hypothesis informed by previous research proposes that the racial composition of NHs, based on the percentage of Black residents, contributes to individual- and facility-level variation in vaccination coverage.[2, 25, 26] Then, the proportion of non-White beneficiaries would increase in NHs with more MA enrollees lowering vaccination rates owing to disparities in care. In this context, we aimed to determine how measures of influenza vaccination offer, receipt and refusal differ among NHs with varying concentrations of residents enrolled in MA.

## METHODS

### Study Design and Data Sources

We conducted a national retrospective cohort study of 100% older adult Medicare beneficiaries residing in NHs during the 2014-2015 influenza season (October 1, 2014-March 31, 2015). We selected this period to identify the study population because it overlaps with the period over which influenza vaccination is entered on the Minimum Data Set (MDS) when received from October 1-March 31. To maximize generalizability, we included all free-standing NHs in the 50 U.S. states, District of Columbia, and Puerto Rico, excluding hospital-based facilities because of significant case-mix and structural differences.[27] We analyzed long-stay (≥100 days) and short-stay (<100 days) NH residents who were ≥65 years of age. A separate analysis of long-stay and short-stay residents was warranted because prior research found differences in risk factors for influenza infection and outcomes between these groups.[28-30] Furthermore, there are potential differences in how reliably influenza vaccination status is captured in the MDS depending on the duration of their NH stay. We used Medicare enrollment data combined with MDS version 3.0 clinical assessments, and facility level data. We obtained NH organizational and aggregate resident characteristics from Certification and Survey Provider Enhanced Reports (CASPER) and LTCFocus.org data. We used CMS’s Nursing Home Compare for overall and domain-specific (staffing, quality, inspections) star rating data.

### NH MA Concentration

We determined a beneficiary’s status of MA coverage at the time of NH admission using Medicare enrollment data. We calculated the percentage of residents in each NH who were enrolled in MA. We used the rank procedure to create a dummy variable categorizing NHs into tertiles (low, moderate, high) based on their percentage of MA enrollees.

### Outcomes

We used the MDS to ascertain vaccination status and reasons for nonreceipt. Although the study population included residents in a NH between October 1st - March 31st, MDS assessments can be submitted with influenza vaccination during those dates through June 30. Therefore, in line with the Nursing Home Compare influenza vaccination quality measures, we assessed all MDS assessments for eligible residents from October 1, 2014 to June 30, 2015.[31] To reduce misclassification of vaccination status we counted beneficiaries who received an influenza vaccine outside the NH during the current influenza season as vaccinated. This consideration is particularly relevant for short-stay residents who are more likely to be vaccinated in the hospital or elsewhere compared with long-stay NH residents. We examined three vaccination measures at the NH level: 1) percentage of residents assessed and appropriately provided the seasonal influenza vaccine; 2) percentage of receipt of influenza vaccine; and 3) percentage of refusal of influenza vaccine.

### Covariates

Our analysis accounted for NH variables that capture the demographic (age, sex, race/ethnicity) composition of residents and their physical and clinical attributes (e.g., acuity index, activities of daily living scale, cognitive function scale) as well as facility structural (e.g., for-profit ownership, bed count, occupancy rates, rurality, payer mix) and quality (overall star rating) characteristics. These were selected based on prior literature and substantive knowledge. The overall star rating is a composite score (ranging from 1 to 5) that takes into account a NH’s performance on staffing, health inspections, and care quality measures. We included the Herfindahl-Hirschman index, which measures the concentration of NH beds in a county, as a covariate to account for variation in NH availability. Additionally, we controlled for the county-level MA penetration rate since MA markets vary substantially. MA penetration is defined as the share of Medicare beneficiaries enrolled in MA plans per county. We used MA penetration data from September 2014 which is the month prior to the start of our observation period.[32] We imputed the state average MA penetration rate for counties with missing or suppressed penetration values due to small sample sizes.

### Statistical Analysis

We compare the characteristics of NHs with different concentrations of residents with MA coverage. To assess the relationship between MA concentration and influenza vaccination rates, we conducted a linear regression analysis specified to account for clustering of residents within facilities and facilities within counties using the Huber-White sandwich estimator via generalized estimating equations. We specified an unstructured working correlation structure. In the model we included the dummy variable for the NH’s MA concentration and the above-described covariates. Variables with a p-value <0.05 were considered associated with the outcome.

### Stability Analysis

We carried out a stability analysis to determine the robustness of the results by applying an alternate and stricter definition for MA enrollment that required MA coverage during the entire observation period instead of only at admission. This analysis provides information on the extent to which switching from MA to FFS after admission affects the results.

### Software, Data Use Agreement, and Ethics Approval

Data preparation and analyses were conducted using SAS version 9.4 (SAS Institute, Inc., Cary, NC). The Brown University Institutional Review Board approved this study.

## RESULTS

### Long-Stay Cohort

From a national total cohort of 1,690,642 Medicare beneficiaries ≥65 years of age, we identified 936,513 long-stay residents living in 12,384 unique Medicare-certified NHs between October 1, 2014 and March 31, 2015 (Table 1). At the resident level, the overall prevalence of MA enrollment at the time of NH admission was 21.4% among long-stay residents. When NHs were classified into three groups by their prevalence of residents enrolled in MA, the groups were classified at the following thresholds: low MA concentration (0% to 3.3%), moderate MA concentration (3.4% to 18.6%), and high MA concentration (>18.6%).

**Table 1:** Resident and nursing home characteristics by category of Medicare Advantage concentration, long-stay residents, 2014-2015

|  |  | Nursing Home Prevalence of Residents with Medicare Advantage |  |  |
| --- | --- | --- | --- | --- |
|  | Overall | 0% - 3.3% | 3.4% - 18.6% | >18.6% |
| **Resident Characteristics** |  |  |  |  |
| Number of individuals | 936,513 | 267,924 | 326,106 | 236,782 |
| Age in years, mean (sd) | 84.0 (8.7) | 83.8 (8.7) | 84.0 (8.7) | 84.2 (8.6) |
| Age category, % |  |  |  |  |
| 65-74 | 18.1 | 18.6 | 18.1 | 17.6 |
| 75-84 | 13.5 | 13.7 | 13.5 | 13.3 |
| 85+ | 68.5 | 67.8 | 68.3 | 69.1 |
| Female sex, % | 70.6 | 69.8 | 70.7 | 71.1 |
| Race/ethnicity, % |  |  |  |  |
| White | 82.8 | 83.4 | 83.8 | 81.5 |
| Black | 12.3 | 12.4 | 11.8 | 12.6 |
| Hispanic | 2.0 | 2.0 | 2.0 | 2.1 |
| Asian | 1.5 | 0.8 | 1.2 | 2.2 |
| Enrolled in MA at NH admission | 21.4 | 10.9 | 10.4 | 40.0 |
| Enrolled in MA during entire study period | 18.2 | 9.4 | 8.7 | 34.3 |
| **Nursing Home Characteristics from OSCAR and LTCFocus** |  |  |  |  |
| Number of facilities | 12,384 | 4,131 | 4,127 | 4,126 |
| MA enrollment at admission, mean (sd) % | 16.8 (19.5) | 0.4 (0.9) | 10.4 (4.3) | 39.6 (17.0) |
| median (Q1, Q3) | 10.2 (0, 25.0) | 0 (0, 0) | 10.2 (6.7, 13.8) | 71.4 (64.7, 77.3) |
| Rural setting, % | 26.9 | 38.4 | 29.3 | 13.1 |
| For profit, % | 73.0 | 70.4 | 75.6 | 73.0 |
| Multifacility/chain, % | 57.8 | 51.4 | 60.4 | 61.7 |
| Total beds, mean (sd) % | 113 (61) | 82 (8) | 116 (56) | 124 (69) |
| Occupancy rate, mean (sd) % | 83.1 (13.8) | 81.4 (14.8) | 83.2 (13.6) | 84.7 (12.6) |
| Black resident composition including non-Medicare, mean (sd) % | 11.6 (18.0) | 11.9 (18.8) | 11.3 (17.6) | 11.6 (17.6) |
| Percent paying with Medicaid, mean (sd) % | 60.1 (22.2) | 62.0 (22.4) | 59.3 (22.6) | 58.9 (21.3) |
| Resident acuity index | 12.2 (1.3) | 11.9 (1.5) | 12.3 (1.2) | 12.3 (1.3) |
| Mean ADL score for residents in facility | 16.6 (2.7) | 15.7 (3.2) | 16.8 (2.4) | 17.1 (2.3) |
| Residents with CHF, mean (sd) % | 20.1 (9.6) | 19.8 (10.4) | 20.2 (9.1) | 20.3 (9.2) |
| Residents with schizophrenia and/or bipolar disorder, mean (sd) % | 9.9 (12.4) | 11.9 (16.1) | 9.0 (10.1) | 8.8 (9.8) |
| Residents with low cognitive function score, mean (sd) % | 33.1 (13.6) | 31.8 (14.8) | 33.0 (13.0) | 34.4 (12.7) |
| Low resource facility based on payer mix, % | 6.66 | 9.1 | 5.7 | 5.2 |
| Herfindal index of nursing home beds in county, mean (sd) | 0.19 (0.23) | 0.24 (0.26) | 0.21 (0.25) | 0.12 (0.17) |
| MA penetration in NH county, mean (sd) % | 28.9 (14.1) | 23.0 (13.2) | 25.2 (11.2) | 38.5 (12.4) |
| Above average MA penetration in county, % | 48.1 | 31.9 | 34.1 | 78.1 |
| **Nursing Home Quality Characteristics** |  |  |  |  |
| High overall Nursing Home Compare rating, 4-5 stars, % | 64.9 | 89.2 | 52.9 | 52.6 |
| Overall star rating (range 1-5 stars), mean (sd) | 3.8 (1.3) | 4.6 (0.9) | 3.4 (1.3) | 3.4 (1.3) |
| Staffing rating, mean (sd) | 3.7 (1.2) | 4.6 (0.9) | 3.3 (1.1) | 3.3 (1.1) |
| Quality rating, mean (sd) | 4.4 (0.9) | 4.8 (0.6) | 4.2 (0.9) | 4.3 (0.9) |
| Inspection/survey rating, mean (sd) | 3.1 (1.2) | 3.7 (0.8) | 2.8 (1.3) | 2.7 (1.3) |

NH: nursing home, SD: standard deviation, MA: Medicare Advantage, ADL: Activities of Daily Living – on a scale of 0 to 28, higher scores indicate more limitations in these activities, CHF: congestive heart failure, CFS: cognitive function score – lower scores indicate greater cognitive impairment
\*The Herfindal index is a measure of nursing home concentration, it ranges from 0 to 1

### Resident and NH Characteristics by MA Concentration

Resident and facility characteristics varied by the prevalence of MA beneficiaries in NHs. As the prevalence of MA-enrolled residents increased, the beneficiaries tended to be older in age, and more racially and ethnically diverse. NHs with the highest prevalence of MA enrollees were more often larger, part of a chain system, and located in urban settings. The resident acuity index varied minimally across categories of MA prevalence. However, NHs with increasing MA prevalence had residents with more limitations in activities of daily living, greater cognitive impairment, but lower levels of serious mental illness than NHs with lower MA prevalence. The majority (89.2%) of NHs with low MA prevalence had a high overall star rating of 4 or 5 compared with about half of NHs in the other MA categories who met the same ratings. The prevalence of MA in NHs was higher for facilities located in counties with greater MA penetration rates.

### Influenza Vaccination Rates by MA Concentration

On average, 96.9% of long-stay residents in NHs with a low prevalence of MA-enrolled residents were assessed and appropriately provided influenza vaccines compared with 94.7% of residents in NHs with the highest prevalence of MA. While rates of influenza vaccine receipt were similar, vaccine refusal decreased as the prevalence of MA enrollees in a NH increased (Figure 1).

**Figure 1:**
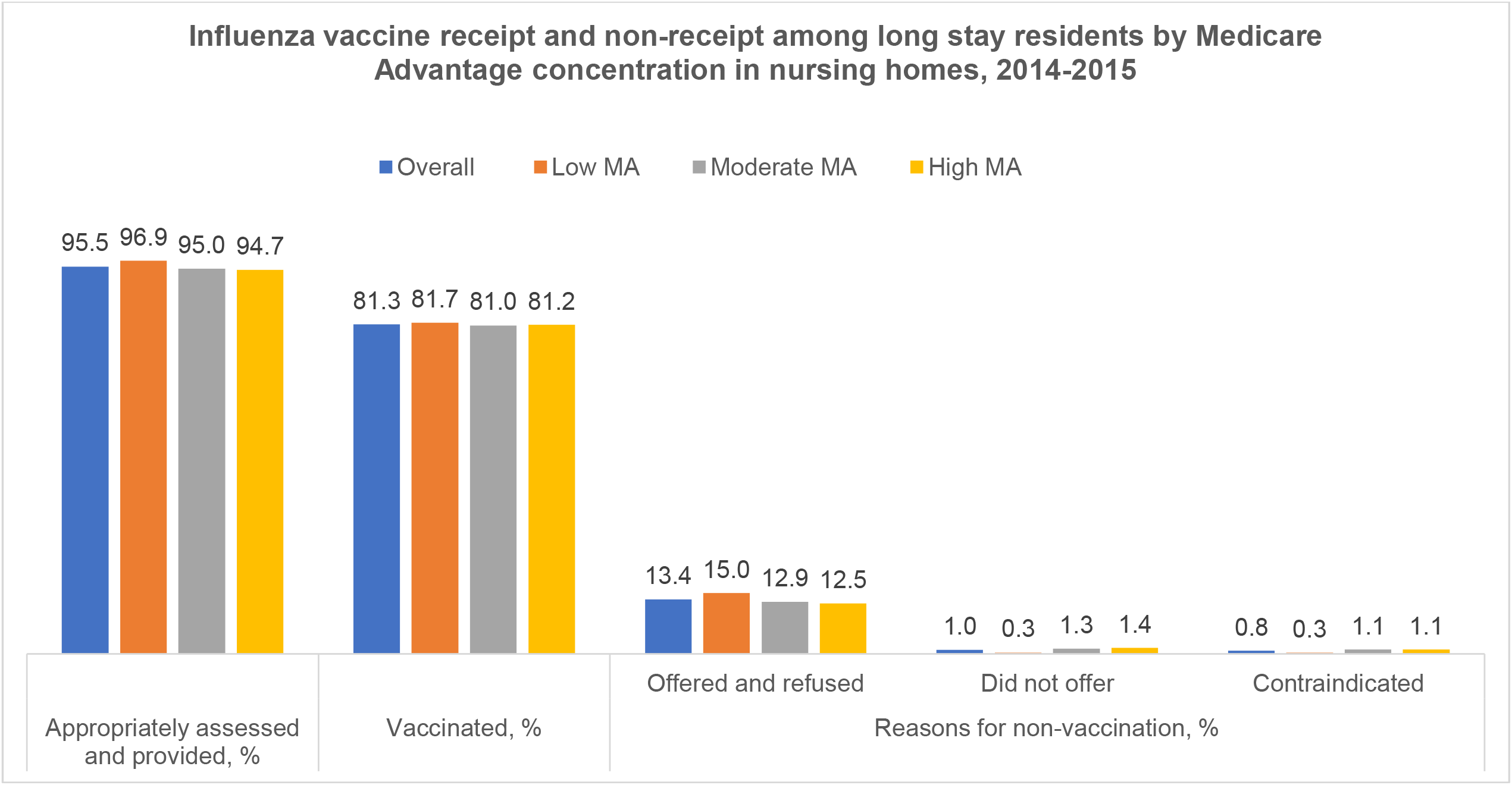
Unadjusted vaccination receipt and non-receipt by MA concentration among long-stay residents, 2014-2015

Table 2 presents the unadjusted and adjusted results from multivariable regression models. The adjusted association between MA concentration and influenza vaccine assessment and provision was minimal in magnitude but statistically significant: low MA (reference) versus high MA (prevalence difference -0.82%, 95% confidence limits [-1.22%, -0.42%] *P*<0.0001).

**Table 2:** Association between NH MA concentration and influenza vaccination rates among long-stay residents, 2014-2015

|  | Appropriately assessed and provided |  | Vaccinated |  | Offered and refused vaccine |  |
| --- | --- | --- | --- | --- | --- | --- |
|  | Prevalence difference |  | Prevalence difference |  | Prevalence difference |  |
|  | Unadjusted, %<br>(95% CL) | Adjusted, %<br>(95% CL) | Unadjusted, %<br>(95% CL) | Adjusted, %<br>(95% CL) | Unadjusted, %<br>(95% CL) | Adjusted, %<br>(95% CL) |
| Low MA Prevalence<br>(n=4,131 NHs; 267,924 people) | reference | reference | reference | reference | reference | reference |
| Moderate MA Prevalence<br>(n= 4,127 NHs; 326,106 people) | -1.97<br>(-2.23, -1.71) | -0.70<br>(-1.06, -0.34) | -0.66<br>(-1.12, -0.21) | 1.70<br>(1.15, 2.24) | -2.11<br>(-2.47, -1.75) | -3.10<br>(-3.53, -2.68) |
| High MA Prevalence<br>(n=4,126 NHs; 236,782 people) | -2.25<br>(-2.52, -1.98) | -0.82<br>(-1.22, -0.42) | -0.54<br>(-0.99, -0.08) | 3.05<br>(2.45, 3.66) | -2.51<br>(-2.87, -2.16) | -4.63<br>(-5.11, -4.15) |

CL: confidence limits

Influenza vaccination rates among long-stay residents were higher in NHs with moderate (1.70%, [1.15%, 2.24%]), or high (3.05%, [2.45, 3.66]) MA compared with NHs with the lowest prevalence of MA (Table 2). Influenza vaccine refusal was lower in NHs with moderate (−3.10% [-3.53%, -2.68%], or high (−4.63% [-5.11%, -4.15%]) MA compared with NHs with the lowest prevalence of MA. All *P* values were <0.0001.

NH variables that were positively associated with higher rates of appropriate assessment and provision and influenza vaccination included mean age, occupancy rate, high NH quality star rating, percent with serious mental illness, percent paying with Medicaid, and the Herfindahl-Hirschman index. In contrast, for profit and chain ownership and increasing percent of Black residents were associated with decreased assessment and appropriate provision, and influenza vaccination. See supplementary Table S1 for covariate estimates and *P* values.

### Secondary Analysis: Short-Stay Cohort

There were 753,616 short-stay residents distributed across 12,205 unique NHs. The person-level prevalence of MA enrollment at the time of admission to a NH was 32.6% (supplementary Table S2). We found much lower levels of assessment and appropriate provision of influenza vaccines (82.8% vs. 95.5% overall) than among long-stay NH residents (Figure S1). There was greater variation in vaccination rates, ranging from 62.7% to 72.3% across MA categories, in contrast to the narrow range from 81.0% to 81.7% among long-stay residents. One in 5 short-stay residents in NHs with high MA prevalence refused influenza vaccination compared with about 1 in 20 of those in NHs with low MA prevalence. After adjustment, there was no association between MA prevalence and influenza vaccine receipt. However, vaccine refusal was higher as the prevalence of MA increased: high vs low MA, 5.15% [4.08%, 6.23%] (Table 3). See supplementary Table S3 for covariate estimates and *P* values for the short-stay analysis.

**Table 3:** Association between NH MA concentration and influenza vaccination rates among short-stay residents, 2014-2015

|  | Appropriately assessed and provided |  | Vaccinated |  | Offered and refused vaccine |  |
| --- | --- | --- | --- | --- | --- | --- |
|  | Prevalence difference |  | Prevalence difference |  | Prevalence difference |  |
|  | Unadjusted, %<br>(95% CL) | Adjusted, %<br>(95% CL) | Unadjusted, %<br>(95% CL) | Adjusted, %<br>(95% CL) | Unadjusted, %<br>(95% CL) | Adjusted, %<br>(95% CL) |
| Low MA Prevalence<br>(n=4,077 NHs; 149,329 people) | reference | reference | reference | reference | reference | reference |
| Moderate MA Prevalence<br>(n= 4,066 NHs; 286,858 people) | 7.29<br>(6.66, 7.92) | 5.33<br>(4.36, 6.30) | -6.67<br>(-7.46, -5.88) | -0.01<br>(-1.29, 1.10) | 13.17<br>(12.51, 13.82) | 5.23<br>(4.28, 6.19) |
| High MA Prevalence<br>(n=4,062 NHs; 317,429 people) | 5.64<br>(4.98, 6.30) | 4.65<br>(3.58, 5.72) | -9.59<br>(-10.43, -8.75) | -0.69<br>(-2.02, 0.64) | 14.42<br>(13.70, 15.14) | 5.15<br>(4.08, 6.23) |

CL: confidence limits

### Stability Analysis: Alternate MA Enrollment Definition

Changing the definition of MA enrollment yielded substantively similar results to the main analysis. There appeared to be a clearer dose response relationship when MA was defined on the basis of enrollment throughout the entire observation period rather than at the time of admission. See supplementary Table S4.

## DISCUSSION

This study investigated influenza vaccination receipt and nonreceipt among older adults in NHs, and their variation on the basis of the concentration of residents enrolled in MA. Several findings stood out in our analysis. First, although nearly all long-stay residents were appropriately assessed (95.5%), influenza vaccination rates were lower (81.3%) largely due to high refusal rates (13.4%) when the vaccine was offered. Second, although crude estimates were similar, in adjusted models we found that as the concentration of MA enrollees increased so did receipt of influenza vaccination among long-stay residents. Third, two-thirds of short-stay residents had documented receipt of influenza vaccination, and there was no relationship between MA concentration and influenza vaccination among short-stay NH residents. Fourth, rates of influenza vaccine refusal decreased with increasing MA concentration in long-stay residents but increased in short-stay residents.

Our finding that NHs with a greater share of MA enrollees have higher influenza vaccination coverage rates among long-stay residents are consistent with perspectives that MA plans promote preventive care use. Given that nearly all the attention on MA efforts to improve preventive care use has targeted community-dwelling beneficiaries, the extent to which MA plans conduct health promotion efforts in NHs is unknown. Individual MA plans conduct care coordination and health promotion efforts for their beneficiaries with varying rigor and success. As such, MA beneficiaries may not experience these benefits uniformly as MA plans are not created equal.[24] The processes that MA plans have in place for outreach and education for providers and patients in NH settings deserve attention in efforts to increase vaccination rates. The importance of addressing this knowledge gap is magnified by the growing enrollment in MA,[8] expensive costs of post-acute and long-term care,[33] and high risk of morbidity and mortality due to respiratory infections in NH residents and older adults generally.[34, 35] The COVID-19 pandemic adds further imperative to explore levers (e.g., care coordination and initiatives to promote preventive care) at the MA plan level to improve NH influenza vaccination coverage.

While improving uptake of the annual influenza vaccine is a perennial challenge,[2] the imminence of a vaccine for COVID-19 means that it will be even more critical to ensure high vaccination rates among NH residents – a population that has experienced disproportionately high rates of COVID-19 cases and deaths. NH residents have accounted for approximately 25% of the documented deaths due to COVID-19 although less than 0.5% of the total U.S population (∼1.5 million people) live in NHs.[36] Our findings indicate that a notable proportion of NH residents decline the influenza vaccine when offered with more refusals in nursing with higher percentages of Black residents and among short-stay compared with long-stay Medicare beneficiaries. Culturally appropriate education campaigns to raise awareness, counter misinformation, and encourage NH residents to accept influenza and COVID-19 vaccinations are needed to more effectively mitigate the transmission of these infections.

Since the composition of residents in a NH often includes a mix of post-acute short-stay and long-stay residents,[37] effective influenza mitigation strategies should also target improving the assessment and appropriate provision of the vaccine to short-stay residents.[30] This may require NHs to maintain vaccine supplies over a longer period during influenza season. Although discrepancies in data collection to determine vaccination status are likely greater among short-stay than long-stay residents, we found that not being offered the influenza vaccine was more common for short-stay residents than long-stay residents. This presents a low-barrier opportunity for NHs to improve their influenza vaccination performance by extending their efforts to offer and vaccinate short-stay residents. Such targeted efforts could be especially beneficial for NHs with large proportions of short-stay residents. In addition, our results suggest actions to improve overall NH Compare star ratings (targeting 4 or 5 starts) could contribute to better vaccination rates. While the quality domain of star ratings includes NH vaccination coverage, this is unlikely to fully explain the strong independent associations of the overall star rating with vaccination rates in multivariable analyses.

This study has limitations. First, vaccination status may be misclassified, especially for short-stay residents who are less frequently assessed. Second, this is a cohort study focusing on a single influenza season (2014-2015). Nonetheless, the findings provide foundational evidence that point to the relevance of further investigation through longitudinal and more recent data. Third, we relied on resident acuity and comorbidity measures from the CASPER database rather than MDS clinical assessments. However, by using CASPER variables we avoided making assumptions that would be required to handle missing data particularly for short-stay residents who more frequently have missing information on MDS-derived variables. Also, our findings may not generalize to beneficiaries younger than 65 years, residing in the community, or with insurance coverage other than Medicare.

In conclusion, this study found that higher concentration of MA beneficiaries in NHs is associated with increased rates of influenza vaccination receipt among long-stay residents. There was no evidence that MA concentration affected influenza vaccination rates among short-stay residents. Vaccine refusal when offered was lower (among long-stay) and greater (among short-stay) as the prevalence of MA beneficiaries increased. As the MA program continues to grow and more MA-enrolled beneficiaries enter NHs, concerted efforts by MA plans and NHs will be essential to improve influenza vaccination rates and reduce vaccine refusals. This importance is magnified in the COVID-19 era when mitigating the transmission of respiratory infections is of critical importance for the health of NH residents and staff.

## Data Availability

The data used in this study are not available for sharing due to restrictions of data use agreements with the U.S. Centers for Medicare and Medicaid Services.

