## Supplemental tables for "Variation in influenza vaccine assessment, receipt, and refusal by the concentration of Medicare Advantage enrollees in U.S. nursing homes"

**SUPPLEMENTARY MATERIALS**

**Table S1:** Estimates from multivariable regression models of vaccination rates among long-stay NH residents, 2014-2015

|  | Assessed and appropriately provided | | Vaccinated | | Offered and refused | |
| --- | --- | --- | --- | --- | --- | --- |
|  | Prevalence difference (%) | *P* value | Prevalence difference (%) | *P* value | Prevalence difference (%) | *P* value |
| MA Concentration Category |  |  |  |  |  |  |
| Low MA | reference | reference | reference | reference | reference | reference |
| Moderate MA | -0.700 | 0.0001 | 1.696 | <0.0001 | -3.103 | <0.0001 |
| High MA | -0.822 | <0.0001 | 3.054 | <0.0001 | -4.629 | <0.0001 |
| Mean age | 0.294 | <0.0001 | 0.948 | <0.0001 | -0.625 | <0.0001 |
| Percent female | 0.014 | 0.40 | -0.048 | 0.0123 | 0.063 | <0.0001 |
| Urban vs. rural setting | -0.486 | 0.0003 | -0.324 | 0.1494 | -0.183 | 0.2998 |
| For profit vs. non-profit | -0.491 | <0.0001 | -2.841 | <0.0001 | 2.379 | <0.0001 |
| Multifacility/chain vs. non-chain | -0.602 | <0.0001 | -1.177 | <0.0001 | 0.562 | 0.0002 |
| Total beds, mean n | 0.001 | 0.1316 | -0.002 | 0.324 | 0.003 | 0.0204 |
| Occupancy rate, mean % | 0.024 | <0.0001 | 0.043 | <0.0001 | -0.021 | 0.0009 |
| Black resident composition including non-Medicare, mean % | -0.034 | <0.0001 | -0.100 | <0.0001 | 0.071 | <0.0001 |
| Percent paying with Medicaid, mean % | 0.037 | <0.0001 | 0.072 | <0.0001 | -0.036 | <0.0001 |
| Resident acuity index | 0.201 | 0.0942 | 0.251 | 0.1303 | -0.085 | 0.4461 |
| Mean ADL score for residents in facility | -0.031 | 0.5256 | -0.116 | 0.0982 | 0.093 | 0.0488 |
| Percent of residents with CHF | 0.006 | 0.3684 | -0.005 | 0.6686 | 0.007 | 0.4752 |
| Percent of residents with schizophrenia and/or bipolar disorder | 0.034 | <0.0001 | 0.048 | <0.0001 | -0.011 | 0.2364 |
| Percent of residents with low cognitive function score, mean | -0.005 | 0.3684 | -0.066 | <0.0001 | 0.057 | <0.0001 |
| Low resource vs. high resource facility based on payer mix | 0.078 | 0.7598 | 0.385 | 0.3889 | -0.315 | 0.3823 |
| Herfindal index for nursing home beds in county, range 0-1 | 0.880 | 0.0001 | 2.453 | <0.0001 | -1.764 | <0.0001 |
| Overall Nursing Home Compare rating of 4-5 stars vs. 1-3 stars | 1.561 | <0.0001 | 2.028 | <0.0001 | -0.400 | 0.038 |
| MA penetration in NH county (above vs. below national average) | -0.019 | 0.8911 | -2.035 | <0.0001 | 2.100 | <0.0001 |

NH: nursing home, SD: MA: Medicare Advantage, ADL: Activities of Daily Living – on a scale of 0 to 28, higher scores indicate more limitations in these activities, CHF: congestive heart failure, CFS: cognitive function score – lower scores indicate greater cognitive impairment

*The Herfindal index is a measure of nursing home concentration, it ranges from 0 to 1

**Table S2:** Resident and Facility characteristics by category of Medicare Advantage concentration: Short-stay residents, 2014-2015

|  |  | Nursing Home Prevalence of Residents with Medicare Advantage (MA) | | |
| --- | --- | --- | --- | --- |
|  | Overall | 0% - 6.2% | 6.3% - 31.3% | >31.3% |
| **Resident Characteristics** |  |  |  |  |
| Number of individuals | 753,616 | 149,329 | 286,858 | 317,429 |
| Age in years, mean (SD) | 80.5 (8.2) | 80.7 (8.2) | 80.6 (8.2) | 80.2 (8.2) |
| Age category, % |  |  |  |  |
| 65-74 | 18.1 | 27.6 | 28.5 | 30.0 |
| 75-84 | 13.5 | 18.1 | 18.1 | 18.4 |
| 85+ | 68.5 | 54.3 | 53.3 | 51.6 |
| Female sex, % | 70.6 | 69.8 | 70.7 | 64.4 |
| Race/ethnicity, % |  |  |  |  |
| White | 86.3 | 88.3 | 88.3 | 83.6 |
| Black | 9.1 | 8.1 | 8.1 | 10.5 |
| Hispanic | 1.7 | 1.4 | 1.1 | 2.4 |
| Asian | 1.3 | 0.8 | 1.1 | 1.6 |
| Residents enrolled in MA at nursing home admission | 32.6 | 17.6 | 19.0 | 51.9 |
| Residents enrolled in MA during entire observation period | 29.9 | 16.0 | 17.0 | 48.1 |
| **Nursing Home Characteristics from OSCAR and LTCFocus** | |  |  |  |
| Number of facilities | 12,205 | 4,077 | 4,066 | 4,062 |
| MA enrollment at admission, mean (sd) % | 23.4 (22.9) | 0.60 (1.5) | 18.8 (7.0) | 51.0 (14.7) |
| median (Q1, Q3) | 18.9 (0, 38.8) | 0 (0, 0) | 18.9 (12.9, 25.0) | 48.9 (38.8, 60.4) |
| Rural setting, % | 26.7 | 38.1 | 27.7 | 14.4 |
| For profit, % | 73.3 | 69.0 | 74.5 | 76.4 |
| Multifacility/chain, % | 58.4 | 52.1 | 59.9 | 63.2 |
| Total beds, mean % (sd) | 113 (61) | 96 (51) | 118 (59) | 125 (67) |
| Occupancy rate, mean (sd) | 82.9 (13.8) | 81.0 (14.9) | 83.3 (13.4) | 84.5 (12.7) |
| Black resident composition including non-Medicare, mean (sd) % | 11.6 (17.9) | 10.7 (17.9) | 10.8 (16.9) | 13.2 (18.9) |
| Percent paying with Medicaid, mean % (sd) | 59.5 (22.2) | 60.6 (22.8) | 57.1 (23.3) | 60.7 (20.4) |
| Resident acuity index | 12.2 (1.3) | 11.9 (1.4) | 12.3 (1.2) | 12.3 (1.3) |
| Mean ADL score for residents in facility | 16.6 (2.6) | 15.8 (3.0) | 17.0 (2.2) | 17.0 (2.3) |
| Percent of residents with CHF, mean % (sd) | 20.3 (9.6) | 20.0 (10.1) | 20.7 (9.4) | 20.1 (9.2) |
| Percent of residents with schizophrenia and/or bipolar disorder | 9.5 (11.3) | 11.1 (14.5) | 8.1 (8.6) | 9.3 (9.8) |
| Percent of residents with low cognitive function score, mean | 33.2 (13.7) | 31.3 (14.3) | 34.0 (13.4) | 34.3 (13.1) |
| Low resource facility based on payer mix, % | 5.8 | 8.0 | 3.9 | 5.6 |
| Herfindal index for nursing home beds in county, range 0-1 | 0.19 (0.23) | 0.24 (0.26) | 0.20 (0.24) | 0.13 (0.18) |
| MA penetration in NH county, mean (sd) % | 28.9 (14.1) | 23.0 (13.2) | 25.2 (11.2) | 38.5 (12.4) |
| Above average MA penetration in NH county, % | 48.2 | 31.0 | 33.3 | 80.3 |
| **Nursing Home Quality Characteristics** |  |  |  |  |
| High overall Nursing Home Compare rating, 4-5 stars, % | 64.6 | 87.2 | 55.2 | 51.4 |
| Overall star rating (range 1-5 stars), mean (sd) | 3.8 (1.3) | 4.6 (0.9) | 3.5 (1.3) | 3.4 (1.3) |
| Staffing rating, mean (sd) | 3.7 (1.2) | 4.6 (0.9) | 3.4 (1.1) | 3.3 (1.1) |
| Quality rating, mean (sd) | 4.4 (0.9) | 4.8 (0.6) | 4.2 (0.9) | 4.3 (0.9) |
| Inspection/survey rating, mean (sd) | 3.1 (1.2) | 3.7 (0.8) | 2.8 (1.3) | 2.7 (1.3) |

NH: nursing home, SD: MA: Medicare Advantage, ADL: Activities of Daily Living – on a scale of 0 to 28, higher scores indicate more limitations in these activities, CHF: congestive heart failure, CFS: cognitive function score – lower scores indicate greater cognitive impairment

*The Herfindal index is a measure of nursing home concentration, it ranges from 0 to 1

**Table S3:** Estimates from multivariable regression models of vaccination rates among short-stay NH residents, 2014-2015

|  | Assessed and appropriately provided | | Vaccinated | | Offered and refused | |
| --- | --- | --- | --- | --- | --- | --- |
|  | Prevalence difference (%) | *P* value | Prevalence difference (%) | *P* value | Prevalence difference (%) | *P* value |
| MA Concentration Category |  |  |  |  |  |  |
| Low MA | reference | reference | reference | reference | reference | reference |
| Moderate MA | 5.328 | <0.0001 | -0.096 | 0.8747 | 5.235 | <0.0001 |
| High MA | 4.650 | <0.0001 | -0.688 | 0.3109 | 5.153 | <0.0001 |
| Mean age | 0.217 | 0.0123 | 1.147 | <0.0001 | -0.886 | <0.0001 |
| Percent female | -0.090 | <0.0001 | 0.064 | 0.0002 | -0.136 | <0.0001 |
| Urban vs. rural setting | 0.392 | 0.2970 | 0.271 | 0.5512 | 0.106 | 0.7720 |
| For profit vs. non-profit | -0.828 | 0.0089 | -3.702 | <0.0001 | 2.857 | <0.0001 |
| Multifacility/chain vs. non-chain | -1.054 | 0.0002 | -1.802 | <0.0001 | 0.743 | 0.0101 |
| Total beds, mean n | 0.003 | 0.1685 | -0.0002 | 0.9541 | 0.004 | 0.0809 |
| Occupancy rate, mean % | 0.048 | <0.0001 | 0.037 | 0.0036 | 0.010 | 0.3609 |
| Black resident composition including non-Medicare, mean % | -0.097 | <0.0001 | -0.162 | <0.0001 | 0.069 | <0.0001 |
| Percent paying with Medicaid, mean % | -0.050 | <0.0001 | -0.003 | 0.7541 | -0.049 | <0.0001 |
| Resident acuity index | -0.180 | 0.2175 | -0.520 | 0.0054 | 0.298 | 0.0629 |
| Mean ADL score for residents in facility | 0.624 | <0.0001 | -0.116 | 0.1639 | 0.439 | <0.0001 |
| Percent of residents with CHF | 0.033 | 0.0223 | 0.063 | 0.0009 | -0.034 | 0.0342 |
| Percent of residents with schizophrenia and/or bipolar disorder | -0.053 | 0.0024 | 0.009 | 0.6728 | -0.065 | 0.0004 |
| Percent of residents with low cognitive function score, mean | 0.095 | <0.0001 | 0.024 | 0.1033 | 0.072 | <0.0001 |
| Low resource vs. high resource facility based on payer mix | 0.882 | 0.2772 | 0.949 | 0.311 | -0.209 | 0.7882 |
| Herfindal index for nursing home beds in county, range 0-1 | 2.674 | <0.0001 | 3.567 | <0.0001 | -0.933 | 0.1757 |
| Overall Nursing Home Compare rating of 4-5 stars vs. 1-3 stars | 2.566 | <0.0001 | 4.031 | <0.0001 | -1.371 | 0.0003 |
| MA penetration in NH county (above vs. below national average) | -0.690 | 0.0339 | -1.928 | <0.0001 | 1.265 | 0.0001 |

NH: nursing home, SD: MA: Medicare Advantage, ADL: Activities of Daily Living – on a scale of 0 to 28, higher scores indicate more limitations in these activities, CHF: congestive heart failure, CFS: cognitive function score – lower scores indicate greater cognitive impairment

*The Herfindal index is a measure of nursing home concentration, it ranges from 0 to 1

**Figure S1**: Unadjusted vaccination receipt and non-receipt by MA concentration among short-stay residents, 2014-2015

Influenza vaccine receipt and non-receipt among short stay residents by Medicare Advantage concentration in nursing homes, 2014-2015

|  | Overall | Low MA | Moderate MA | High MA |
| --- | --- | --- | --- | --- |
| Appropriately assessed and provided, % | 82.7636 | 78.459 | 85.7478 | 84.097 |
| Vaccinated, % | 66.881 | 72.2951 | 65.6276 | 62.7014 |
| Reasons for non-vaccination, % Offered and refused | 14.8852 | 5.69889 | 18.8679 | 20.1188 |
| Reasons for non-vaccination, % Did not offer | 2.80095 | 1.07721 | 3.32758 | 4.00391 |
| Reasons for non-vaccination, % Contraindicated | 0.99747 | 0.465033 | 1.25226 | 1.27683 |

**Table S4:** Stability analysis of the association between NH MA concentration and influenza vaccination rates among long-stay residents. With MA defined based on enrollment throughout the entire observation period

|  | Appropriately assessed and provided | | Vaccinated | | Offered and refused vaccine | |
| --- | --- | --- | --- | --- | --- | --- |
|  | Prevalence difference | | Prevalence difference | | Prevalence difference | |
|  | Unadjusted, % (95% CL) | Adjusted, % (95% CL) | Unadjusted, % (95% CL) | Adjusted, %  (95% CL) | Unadjusted, % (95% CL) | Adjusted, %  (95% CL) |
| Low MA Prevalence  (n=4,131 NHs; 267,924 people) | reference | reference | reference | reference | reference | reference |
| Moderate MA Prevalence  (n= 4,127 NHs; 326,106 people) | -2.16  (-2.42, -1.89) | -0.93  (-1.29, -0.57) | -0.89  (-1.36, -0.42) | 1.37  (0.82, 1.92) | -2.00  (-2.37, -1.63) | -2.93  (-3.36, -2.49) |
| High MA Prevalence  (n=4,126 NHs; 236,782 people) | -2.14  (-2.41, -1.88) | -0.78  (-1.16, -0.40) | -0.0038  (-0.45, 0.44) | 3.28  (2.69, 3.87) | -2.93  (-3.28, -2.59) | -4.79  (-5.25, -4.33) |

CL: confidence limits
